# Time-dependent dynamic transmission potential and instantaneous reproduction number of COVID-19 pandemic in India

**DOI:** 10.1101/2020.07.15.20154971

**Authors:** Gurpreet Singh, Seema Patrikar, P Sankara Sarma, Biju Soman

## Abstract

**Introduction:** Dynamic tools and methods to assess the ongoing transmission potential of COVID-19 in India are required. We aim to estimate time-dependent transmissibility of COVID-19 for India using a reproducible framework.

**Methods:** Daily COVID-19 case incidence time series for India and its states was obtained from https://api.covid19india.org/ and pre-processed. Bayesian approach was adopted to quantify transmissibility at a given location and time, as indicated by the instantaneous reproduction number (R_eff_). Analysis was carried out in R version 4.0.2 using *“EpiEstim_2.2-3”* package. Serial interval distribution was estimated using “uncertain_si” algorithm with inputs of mean, standard deviation, minimum and maximum of mean serial interval as 5.1, 1.2, 3.9 and 7.5 days respectively; and mean, standard deviation, minimum, and maximum of standard deviations of serial interval as 3.7, 0.9, 2.3, and 4.7 respectively with 100 simulations and moving average of seven days.

**Results:** A total of 9,07,544 cumulative incident cases till July 13^th^, 2020 were analysed. Daily COVID-19 incidence in the country was seen on the rise; however, transmissibility showed a decline from the initial phases of COVID-19 pandemic in India. The maximum R_eff_ reached at the national level during the study period was 2.57 (sliding week ending April 4^th^, 2020). R_eff_ on July 13^th^, 2020 for India was 1.16 with a range from 0.59 to 2.98 across various states/UTs.

**Conclusion:** R_eff_ provides critical feedback for assessment of transmissibility of COVID-19 and thus is a potential dynamic decision support tool for on-ground public health decision making.

## Introduction

As of July 13^th^, 2020, 12,768,307 confirmed cases of Novel Coronavirus disease (COVID-19) have occurred in the world, which includes 566,654 deaths.^1^ India is the second-most populous country in the world with 17% of the global population.^2^ The cumulative number of COVID-19 cases reported from the country is increasing steadily, and presently approaching one million cumulative cases, India is having the third largest cumulative case burden of COVID-19 in the globe.^3^ The spread and pattern of the disease in India and its control strategies will be a designing factor for the ongoing pandemic in the coming months. The country is still evolving in the pandemic and efforts by the public health authorities, and the government has been appreciable in the pandemic containment; however, we need dynamic tools and methods to assess the ongoing transmission potential of COVID-19 in the country.

The reproduction number of the infectious agent is the most reliable and sensitive summary statistic of the ever-changing transmission potential in an ongoing epidemic for public health decision making.^4^ When calculated at the introduction of an infected person in a totally susceptible population, it defines the number of individuals who will get infected by the infected person over the infectious period.^5^ However, as the population starts getting infected, the effective reproduction number (R_eff_) at a given point in time becomes the guiding force for decision making. R_eff_ of greater than one at a given time suggests that the caseload and epidemic burden of disease is on the increase. R_eff_ of one is said to be present when the incidence of infections in a population becomes equal to the rate of recovery or deaths. Subsequent to the decline of R_eff_ to less than one, a decline in the epidemic curve and incidence rates are observed in populations.^6^ Hence, all the control efforts made by health authorities including Ministry of Health and Family Welfare across countries should be aimed at reducing the effective reproduction number to less than one to curtail an ongoing epidemic or pandemic as in case of COVID-19.

The mathematical models are a powerful tool to guide public health decision making. Model development requires critical inputs and assumptions.^5,6^ As a consequence, determining static, as well as dynamic parameters for modelling novel epidemics and pandemics, requires empirical inputs based on real-time data. Local estimates based on empirical data will help to formulate realistic and context-specific models and also help to guide, monitor and evaluate public health interventions. In the initial stages of an epidemic, it is technically challenging to calculate R_eff_.^7^ However, as we are now more than six months into the COVID-19 pandemic, R_eff_ estimates are required as vital decision support tool which can complement and enhance existing Public Health mechanisms in COVID-19 prevention and control.

In such a situation, the estimation of transmissibility of COVID-19 in a given location and time based on R_eff_ is of paramount importance. The decision on hotspot identification, allocation of constrained resources such as health personnel, budgetary allocations, equipment accessibility and availability, and decisions on non-pharmacological interventions can be guided by R_eff_ across geographical locations. Further, the assessment and evaluation of interventions and decisions regarding intensification or relaxation of public health measures can be supported with empirical evidence on context-specific transmissibility status of COVID-19. Despite the fact, to the best of our knowledge, there is no such kind of study from India wherein R_eff_ for the country and states have been calculated.

In the COVID-19 pandemic at present, a dynamic and real-time empirical data-based decision support system/ tool is the need of the hour for the administrators and public health professionals at national, state, and district levels for making evidence-informed decisions. In the present study, we aim to estimate the instantaneous reproduction number for India as well as for the states/UTs across the country.

## Methods

### Data source

Crowdsourced line listing of COVID-19 cases in India, freely available from the site, https://api.covid19india.org/ was downloaded^8^ up to 1005 hours on July 14^th^, 2020. From the raw dataset, details regarding daily lab-confirmed COVID-19 incidence (time series data) were extracted. Data were cleaned and pre-processed into tidy incidence time series for analysis. Data triangulation and checks with data available from the Ministry of Health and Family Welfare, India and respective state’s websites was carried out. Randomly chosen government official data sources and downloaded data were tallied, and minor inconsistencies in the dataset were corrected. Epidemic curves were constructed using daily incidence data.

### Understanding the model

The Bayesian approach was adopted to quantify the transmissibility situation at a given point in time, as indicated by R_eff._ The model estimated the average or mean transmissibility of COVID-19 in a given location based on two major parameters. The first parameter was a discrete distribution of serial interval, defined as the time difference between the onset of symptoms in the infector (index) and infectee(secondary) cases. It was assumed that the probability (*p*_*i*_) of an infector transmitting the infection to a susceptible infectee follows gamma distribution and is thus dependent upon the time since infector is infected, but is independent of the stage of pandemic. Thus, an infected person becomes most infectious when *p*_*i*_ is maximum. The second parameter was the number of infected individuals in a location at that time which is indicated by daily COVID-19 incidence data. Hence, at a given time, the infectivity in a given population is dependent upon the number of infective individuals in the population as well as the time since each of them became infected.

### Statistical analysis

All the analysis was carried out in R version 4.0.2 software. The Effective reproduction numbers were estimated using *“EpiEstim_2.2-3”* package. Additional packages used for pre-processing and tidying data included *“lubridate_1.7.9”*, and the metapackage *“tidyverse_1.3.0”*. There are a total of five algorithms in *“EpiEstim_2.2-3”* package for configuring serial interval distributions. Depending upon the available parameters, a specific algorithm needs to be adopted.^7^ The method for calculation of effective reproduction number in the present study was based on “uncertain_si” algorithm for serial interval parameters. Based on literature review (table 1), eight parameters were considered for configuring serial interval distribution:-the mean, standard deviation, minimum and maximum of mean serial intervals were 5.1, 1.2, 3.9 and 7.5 days respectively; and the mean, standard deviation, minimum, and maximum of standard deviations of serial intervals were 3.7, 0.9, 2.3, and 4.7 respectively. The size of the sample for serial interval distributions to be drawn computationally was kept at 100 and a sliding scale of seven days was used for the calculation of R_eff_. Since the study involved publicly available secondary datasets, Institutional Ethics Committee approval was not mandated.

**Table 1.** Review of study characteristics and their key findings used to calculate serial interval distribution for COVID-19 in India.

| S. No | Study | Characteristics | Mean SI (95% CI) | SD SI (95% CI) |
| --- | --- | --- | --- | --- |
| 01 | Nishiura H et al<sup>17</sup> | Estimation of serial interval based on findings from 28 infector-infectee pairs. | Based on all pairs: 4 days (3.1-4.9)<br>Based on 18 most certain pairs: 4.8 days (3.8-6.1) | Based on all pairs: 2.9 days (1.9-4.9)<br>Based on 18 most certain pairs: 2.3 days (1.6-3.5) |
| 02 | Du et al<sup>18</sup> | Observations from 468 transmission events of COVID-19 in mainland China. | Overall findings: 3.9 days (3.5–4.4)<br>In household settings: 4 days (3.1–4.9) | 4.7 days (4.5–5.1) |
| 03 | Ganyani et al<sup>15</sup> | Based on reported contact information of 54 cases from Singapore and 135 cases from Tianjin, China | Singapore: 5.2 days (-3.35 - 13.94)<br>Tianjin (China): 3.95 days (-4.5 - 12.5) | Singapore: 4.32 days (4.1 - 5.6)<br>Tianjin (China): 4.24 days (4.0 - 4.9) |
| 04 | WHO situation report<sup>19</sup> | Estimated Mean SI for COVID-19 | 5-6 days | - |
| 05 | Chan et al<sup>14</sup> | Observation from 47 infector-infectee pairs | 6.5 days | 4.7 days |
| 06 | Li et al<sup>20</sup> | Based on data from 6 infector-infectee pairs | 7.5 days (5.3 - 19) | 3.4 days |
| 07 | Zhao et al<sup>21</sup> | Based on 21 infector-infectee pairs in Hong Kong | 4.4 days (2.9–6.7) | 3.0 days (1.8–5.8) |

## Results

A total of 9,07,544 cumulative incident cases data were analyzed in the present study. The data repository provided daily lab-confirmed incidence data from March 14^th^, 2020 to July 13^th^, 2020: total duration of 122 days.

The epidemic curve, the estimated instantaneous reproduction number and the serial interval distributions for India are represented in figure 1. As shown in the figure, though the incident cases in the country are on the rise at majority of time stamps, the effective reproduction number has shown a decline from the initial phases of the pandemic in India. The maximum instantaneous reproduction reached at the national level during the study period was 2.57 (based on the sliding week ending on April 4^th^, 2020). The state-wise epidemic curves, estimated instantaneous reproduction numbers, and serial interval distributions are given as supplement 1 to the manuscript.

**Figure 1.**
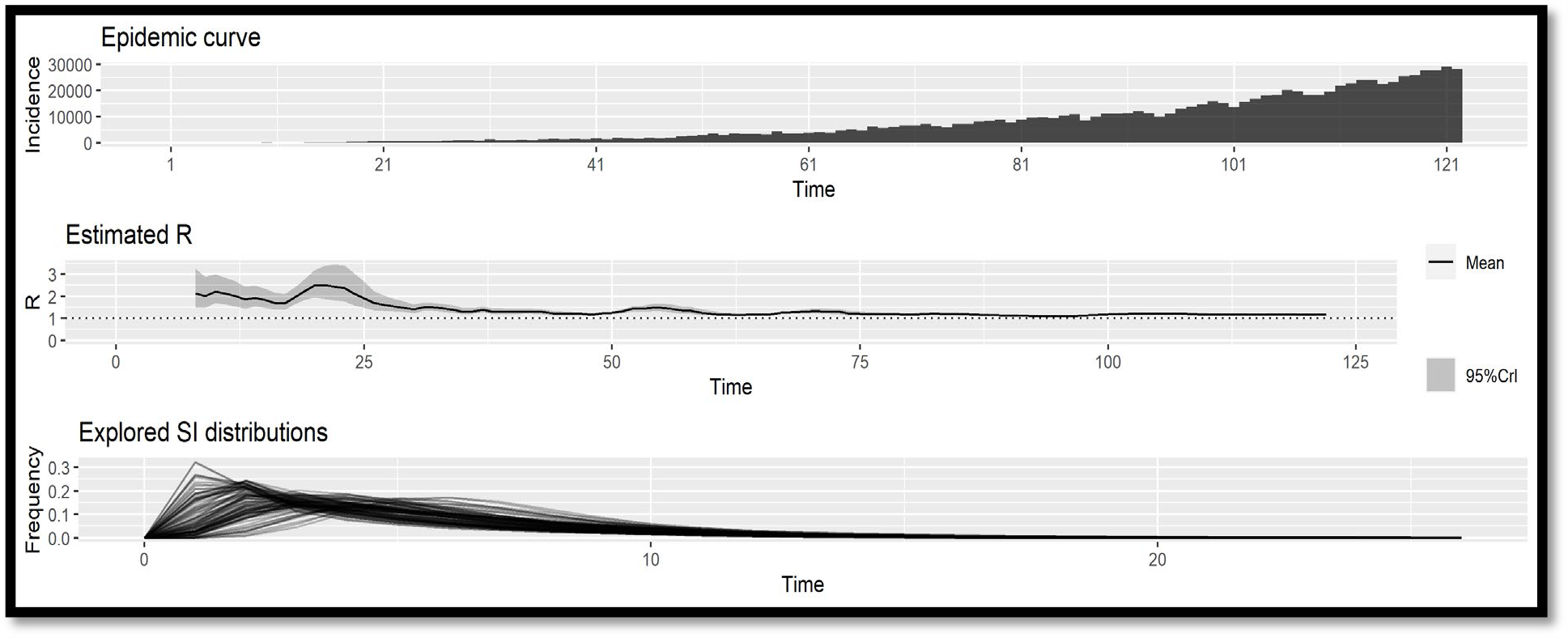
Epidemic curve and effective reproduction number of COVID-19 in India during the study period (March 14^th^ 2020 to July 12^th^ 2020)

Use of estimated instantaneous reproduction number as a support tool for public health decision making is represented in figure 2. The figure represents the overall progression of the estimated instantaneous COVID-19 reproduction numbers for India and its selected states. Figure 2A represents the historical monitoring application of the concept. The estimated instantaneous reproduction number at the national level as well as across the majority of states in the country in the initial stages of the pandemic were higher and have declined on average with the implementation of various public health interventions. Figure 2B represents changes in estimated instantaneous reproduction numbers in the past one month. Since the estimated R_eff_ at national level presently is 1.16, the daily incident COVID-19 cases from the country are still on the rise (as evident from the epidemic curve in figure 1). Among larger states in the country, the state of Delhi has shown a decline of R_eff_ to less than one (figure 2B) in past few weeks, and a decline in the daily incidence of COVID-19 from the state is evident subsequent to the achievement of R_eff_ of less than one (figure S8 Supplement 1). However, as seen in figure 2B, the R_eff_ for the state of Kerala, West Bengal, Karnataka, Punjab, and Maharashtra is larger than one. Further, the slope of R_eff_ is positive for the state of Kerala, West Bengal, Punjab and Tamil Nadu. Day-wise changes in R_eff_ for India at the national level is given as supplement 2A to the manuscript.

**Figure 2:**
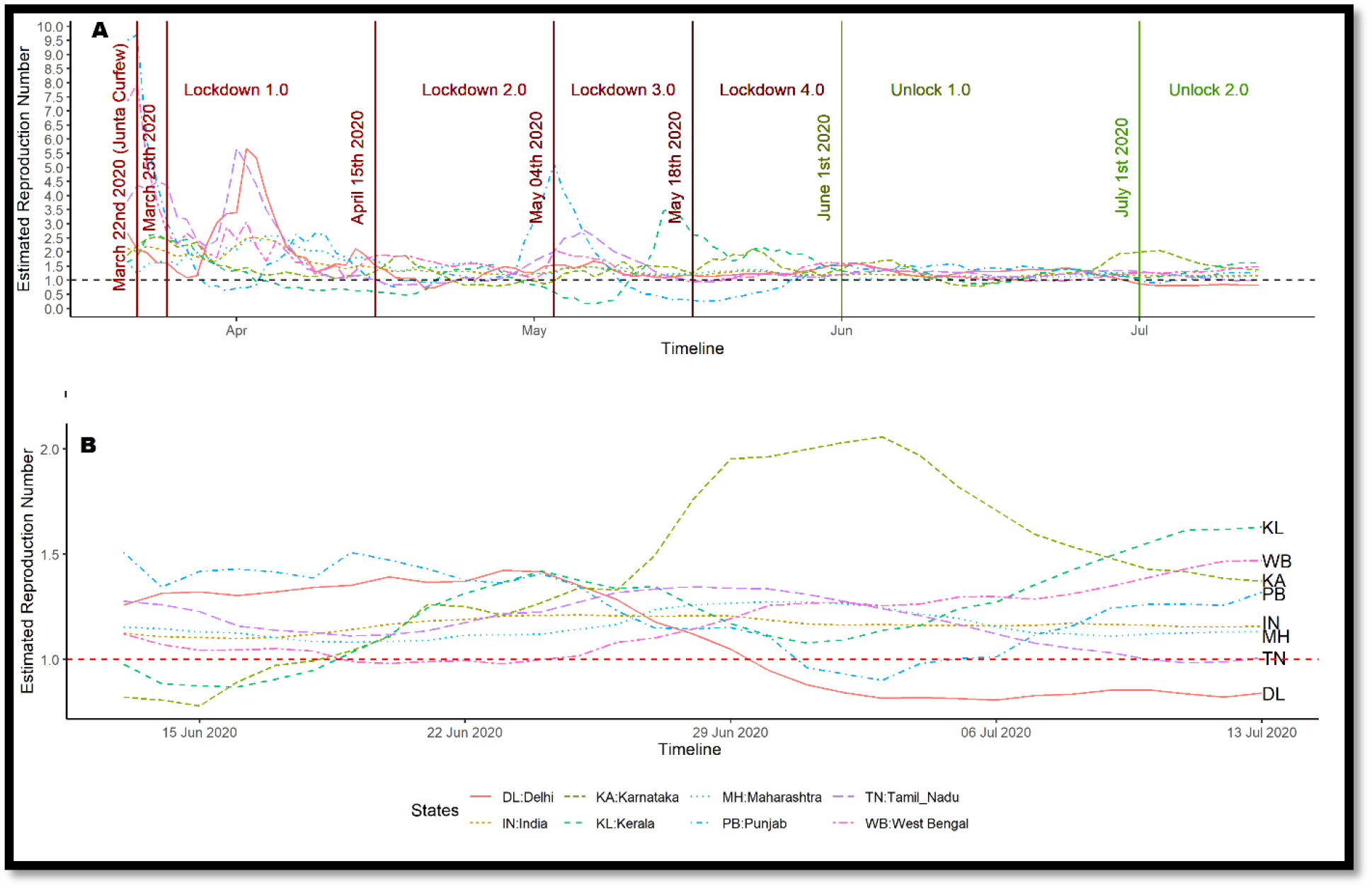
Illustrative example on use of effective reproduction number monitoring as a decision support tool for COVID-19 surveillance. **(A) Effective reproduction number for India and selected high burden states over the pandemic period** **(B) Effective reproduction number in India for the last month**.

**Fig 3:**
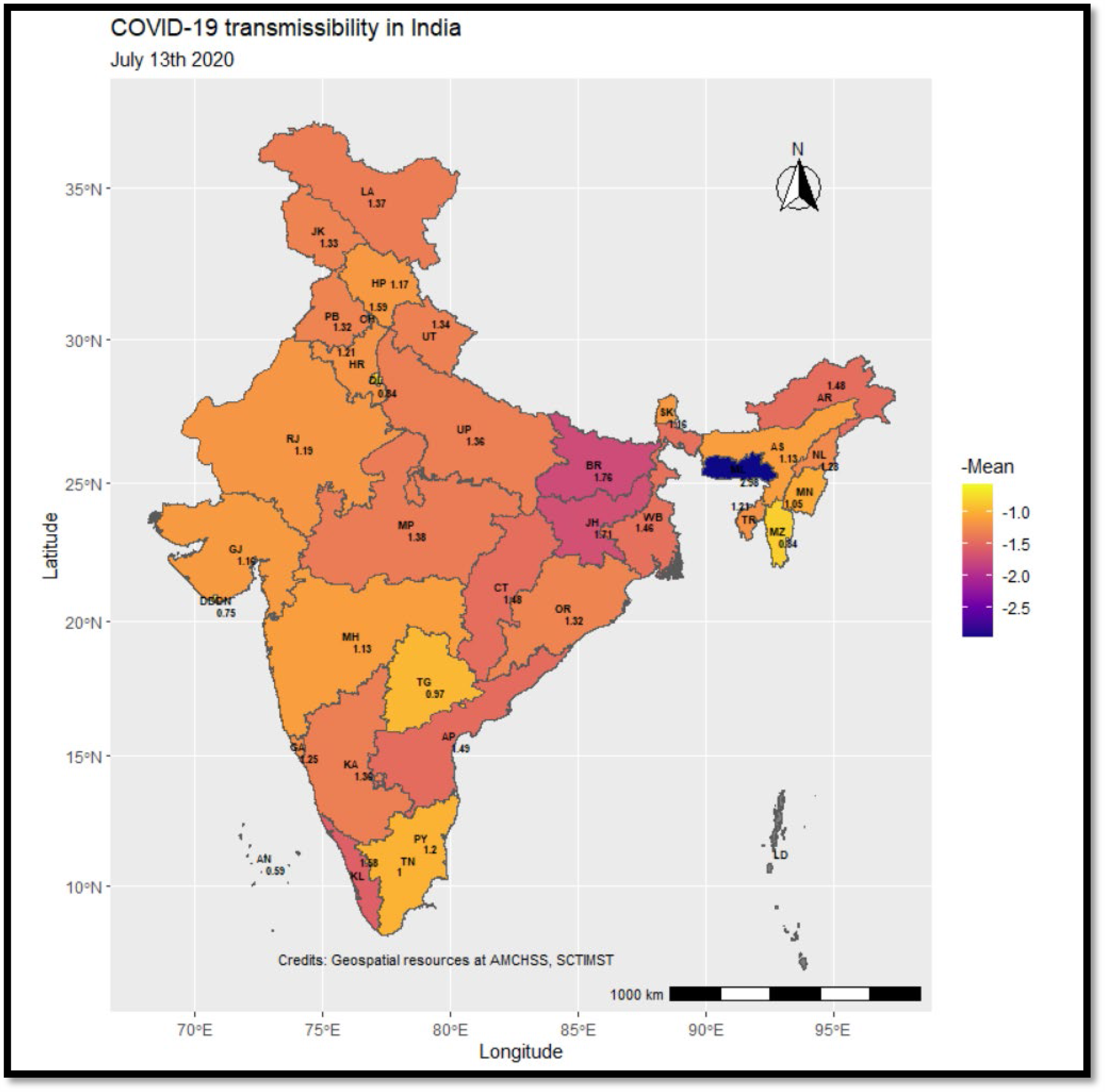
COVID-19 time dependent transmissibility (July 13^th^, 2020) in India. *AN = Andaman and Nicobar Islands, AP = Andhra Pradesh, AR = Arunachal Pradesh, AS = Assam, BR = Bihar, CH = Chandigarh, CT = Chhattisgarh, DDDN = Daman and Diu and Dadar and Nagar Haveli, GA = Goa, GJ = Gujarat, HP = Himachal Pradesh, HR = Haryana, JH = Jharkhand, JK = Jammu and Kashmir, KA = Karnataka, KL = Kerala, LA = Ladakh, MH = Maharashtra, ML = Meghalya, MN = Manipur, MP = Madhya Pradesh, MZ = Mizoram, NL = Nagaland, OR = Odisha, PB = Punjab, PY = Puducherry, RJ = Rajasthan, SK= Sikkim, TG = Telangana, TR = Tripura, UP = Uttar Pradesh and WB = West Bengal.

Mean R_eff_ for India and various states in the country for July 13^th^, 2020 (based on sliding week July 7^th^, 2020 to July 13^th^, 2020) is represented in fig 2. The mean (SD) estimated R_eff_ for India on July 13^th^, 2020 was found to be 1.16 (0.03). Statewide variations were noticed across the states with state wise R_eff_ ranging from 0.59 to 2.98. State-wise mean, first quartile, median and third quartile of the estimated R_eff_ is given in supplement 2B to the manuscript.

## Discussion

The present study documents and demonstrates the use of instantaneous reproduction number in a reproducible framework as public health decision support tool for COVID-19 in India. Instantaneous reproduction number also called the effective reproduction number or time-dependent reproduction number has the potential of strengthening COVID-19 disease surveillance programme in India at the national, state and district levels. Also, being a composite indicator of the transmissibility status in an epidemic situation, R_eff_ provides a critical feedback mechanism for assessment of on-ground public health interventions in real-time settings. Though the present study estimated R_eff_ for the national and state levels, its application at higher spatial granularity in the districts and blocks is vital for understanding COVID-19 transmissibility situation in India.

Reproduction number in a population is based on three major parameters viz. infectivity of a disease agent, duration of infectiousness and the probability of an effective contact resulting in successful transmission of infection.^9^ Biologically, the variations in infectivity and infectiousness is minimal in populations across the globe for a given disease agent, but the probability of effective contact depends heavily on social factors such as population density, socio-economic profile, age structure, gender norms, and individual behaviours among others. Therefore, reproduction numbers vary from place to place. In the present study, we estimated that the maximum mean R_eff_ for India was 2.57 in the initial phase of the pandemic and has reduced to 1.16 at present. Calculation of reproduction numbers for Indian context previously also have documented value of 2.6 (95% CI=2.34, 2.86) in the initial stages and 1.57 (95% CI=1.3, 1.84) for the post lockdown period.^10^

Basic reproduction number of COVID-19 is a static number calculated when the entire population is susceptible. On the other hand, the effective reproduction number is dynamic and natural and is useful for planning purposes. Being instantaneous, it varies with time. In the early stage of spread in India, as shown in figure 2, the R_eff_ was high in certain states as compared to national figures (supplement 1). Till date, the highest effective reproduction number for COVID-19 was reported from the Diamond Princess Cruise Ship epidemic, which was as high as 11. The population mixing pattern and human density in many parts of the country, Dharavi (Maharashtra) as an illustrative example, is likely to be higher than the closed ship settings. Also, very high R_eff_ is difficult to attribute retrospectively to true changes or changes resulting from strengthening of prevention and control measures such as enhanced testing rates, increased awareness, increased reporting efficiency, and modified reporting criteria among others during an ongoing pandemic.^7^ More importantly, such changing trends provide crucial information for action in the ongoing interventions.

Estimation of R_eff_ based on the time of infection and generation time is considered as ideal; however, the data on symptom onset is recorded in routine surveillance since estimating the time of infection is operationally challenging.^5^ As in case of influenza, measles, and SARS, the time of symptom onset in an infected person is temporally close to the onset of infectiousness in COVID-19 also, thus, distribution of generation time and serial interval are theoretically similar. Further, the sliding scale of seven days was chosen in the present study. Depending upon the daily COVID-19 incidence rates, the same can be modified taking posterior probability precision estimates into consideration.^7^ As a result, on the one hand, prompt assessment mechanisms for hotspot containment and control can be developed, and on the other, more extended duration intervention assessments can also be carried out.

The R_eff_ of more than one suggests an increasing trend in the daily incidence of COVID-19 in the present study (figure 1: Epidemic curve), however, sustained check on the rise of R_eff_ at national as well across states substantiates the impact of interventions taken in the country. The impact of the Lockdown implementation in stages is evident from the fact that the effective reproduction number has remained in check and reduced to 1-1.5 in recent weeks. Junta curfew was implemented on March 22^nd^, 2020 followed by nationwide lockdown phase 1, 2, 3 and 4 from March 25^th^ to April 14^th^, April 15^th^ to May 03^rd^, May 4^th^ to May 17^th^, and May 18^th^ to May 31^st^, 2020 respectively. Relaxation of strategies in form of Unlock 1.0 was implemented from June 1^st^ to June 30^th^, 2020 and Unlock 2.0 at present.^11^ However, the economic impact of lockdowns is a concern for national as well as individual state’s perspectives.^12^ Thus, the need for strict lockdown measures, its geographical extent, the timeline of implementation, alternative non-pharmacological interventions, and unlock procedures need assessment with robust epidemiological indicators.

Limitations of the study. The present study was based on crowdsourced publicly available datasets. The data sources for the datasets include multiple official websites by the government of India and is likely to be close to the actual government data, real-time use of National Surveillance Data should be considered for public health decision making.^13^ Further, the present study did not categorise daily COVID-19 incidence between local and imported cases. As a result, it is likely that we might have overestimated the transmissibility of COVID-19 in certain states which have high rates of immigration from expatriates or within-country migration. The overestimation of R_eff_ is justified considering its impact on decision making; however, the provision of modelling categorized cases within the algorithm is available and should be used for enhanced accuracy of the transmissibility estimates. The present study assumed serial interval parameters from the literature. These estimates are likely to be applicable to the Indian settings, health surveillance data from the existing public health programmes needs to be incorporated for developing country-specific decision support system. Further, assessment of the probability distribution function parameters based on empirical data from infector-infectee pairs in the country is required for fine-tuning of the country or state-specific COVID-19 management strategies. In studies on estimating epidemiological parameters in Hongkong, Singapore and Tianjin, gamma distribution of serial interval was found.^14,15^ The effect of serial intervention distribution pattern is theoretically considered to be minimal on effective reproduction number, planning of local public health interventions should be modelled as close to the reality as possible. This requires in-depth studies on estimation of serial interval characteristics from the country at multiple administrative levels. The strength of the chosen algorithm in the present study comes with the proven credibility of “*EpiEstim*” R package. The algorithm’s simulation for outbreaks of historical significance including pandemics was consistent with the available literature and its potential to provide epidemiological insights into pandemics and epidemics resulting from novel diseases has been documented.^7^ Further, taking real-time intervention assessment for COVID-19 as an illustrative example substantiates potential of the algorithm. Thiruvananthapuram district has one of the two large community clusters of COVID-19 identified till now in Kerala.^16^ With the rise in the number of cases, the district was placed under triple lockdown(stringent lockdown) from July 6^th^, 2020 onwards. As shown in SFig.21 in supplement, the number of daily cases are still rising in Kerala, but the slope of R_eff_ for Kerala has started showing a decline (Fig. 2B) suggesting decreased transmissibility of COVID-19 as the impact of the triple lockdown measures. This supports R_eff_ as a near real-time and sensitive indicator for assessment of the epidemic

To conclude, the present study is first of its kind on estimating transmissibility situation of COVID-19 from India. As complementary to existing public health measures for COVID-19, real-time calculations of effective reproduction numbers by national, state and district health authorities should be undertaken as a public health decision support tool. Development of a web-based platform capable of fetching real-time updates in pandemic incidence and using algorithms for estimating transmissibility status on government official servers shall enhance understanding of transmission dynamics of COVID-19 for its prevention and control in India.

## Data Availability

The study is an analysis of secondary data extracted from the publicly available resources (https://api.covid19india.org/). The R codes used for analysis shall be shared with individual researchers on on request.

## Acknowledgement

We acknowledge the services of the proponents and contributors of the crowdsourced line listing of COVID-19 cases in India, https://api.covid19india.org/, which made it possible to undertake this work. We also thank Dr Engelbert Niehaus (Professor for Mathematics and Head of Computer Science Centre at University Koblenz-Landau, Germany) for his valuable guidance.

**<u>Supplementary material 1:</u>** State/UT wise epidemic curves and estimated effective reproduction numbers of COVID-19 in India.

**<u>Supplementary material 2A:</u>** Estimated mean, SD, first quartile, median, and third quartile of instantaneous reproduction numbers for COVID-19 in India

**<u>Supplementary material 2B:</u>** State wise estimated mean, SD, first quartile, median, and third quartile of instantaneous reproduction numbers for COVID-19 on July 13^th^ 2020 in India.

